# Systematic evaluation and external validation of 22 prognostic models among hospitalised adults with COVID-19: An observational cohort study

**DOI:** 10.1101/2020.07.24.20149815

**Authors:** Rishi K. Gupta, Michael Marks, Thomas H. A. Samuels, Akish Luintel, Tommy Rampling, Humayra Chowdhury, Matteo Quartagno, Arjun Nair, Marc Lipman, Ibrahim Abubakar, Maarten van Smeden, Wai Keong Wong, Bryan Williams, Mahdad Noursadeghi on behalf of The UCLH COVID-19 Reporting Group

**Author notes:** Correspondence Prof Mahdad Noursadeghi, Division of Infection & Immunity, Cruciform Building, University College London, London WC1E 6BT, United Kingdom. Members listed in Acknowledgements.

## Abstract

**Background:** The number of proposed prognostic models for COVID-19, which aim to predict disease outcomes, is growing rapidly. It is not known whether any are suitable for widespread clinical implementation. We addressed this question by independent and systematic evaluation of their performance among hospitalised COVID-19 cases.

**Methods:** We conducted an observational cohort study to assess candidate prognostic models, identified through a living systematic review. We included consecutive adults admitted to a secondary care hospital with PCR-confirmed or clinically diagnosed community-acquired COVID-19 (1^st^ February to 30^th^ April 2020). We reconstructed candidate models as per their original descriptions and evaluated performance for their original intended outcomes (clinical deterioration or mortality) and time horizons. We assessed discrimination using the area under the receiver operating characteristic curve (AUROC), and calibration using calibration plots, slopes and calibration-in-the-large. We calculated net benefit compared to the default strategies of treating all and no patients, and against the most discriminating predictor in univariable analyses, based on a limited subset of *a priori* candidates.

**Results:** We tested 22 candidate prognostic models among a cohort of 411 participants, of whom 180 (43.8%) and 115 (28.0%) met the endpoints of clinical deterioration and mortality, respectively. The highest AUROCs were achieved by the NEWS2 score for prediction of deterioration over 24 hours (0.78; 95% CI 0.73-0.83), and a novel model for prediction of deterioration <14 days from admission (0.78; 0.74-0.82). Calibration appeared generally poor for models that used probability outcomes. In univariable analyses, admission oxygen saturation on room air was the strongest predictor of in-hospital deterioration (AUROC 0.76; 0.71-0.81), while age was the strongest predictor of in-hospital mortality (AUROC 0.76; 0.71-0.81). No prognostic model demonstrated consistently higher net benefit than using the most discriminating univariable predictors to stratify treatment, across a range of threshold probabilities.

**Conclusions:** Oxygen saturation on room air and patient age are strong predictors of deterioration and mortality among hospitalised adults with COVID-19, respectively. None of the prognostic models evaluated offer incremental value for patient stratification to these univariable predictors.

## Introduction

Coronavirus disease 2019 (COVID-19), caused by severe acute respiratory syndrome coronavirus-2 (SARS-CoV-2), causes a spectrum of disease ranging from asymptomatic infection to critical illness. Among people admitted to hospital, COVID-19 has reported mortality of 21-33%, with 14-17% requiring admission to high dependency or intensive care units (ICU)^1–4^. Exponential surges in transmission of SARS-CoV-2, coupled with the severity of disease among a subset of those affected, pose major challenges to health services by threatening to overwhelm resource capacity^5^. Rapid and effective triage at the point of presentation to hospital is therefore required to facilitate adequate allocation of resources and to ensure that patients at higher risk of deterioration are managed and monitored appropriately. Importantly, prognostic models may have additional value in patient stratification for emerging drug therapies^6,7^.

As a result, there has been global interest in development of prediction models for COVID-19^8^. These include models aiming to predict a diagnosis of COVID-19, and prognostic models, aiming to predict disease outcomes. At the time of writing, a living systematic review has already catalogued 145 diagnostic or prognostic models for COVID-19^8^. Critical appraisal of these models using quality assessment tools developed specifically for prediction modelling studies suggests that the candidate models are poorly reported, at high risk of bias and over-estimation of their reported performance^8,9^. However, independent evaluation of candidate prognostic models in unselected datasets has been lacking. It therefore remains unclear how well these proposed models perform in practice, or whether any are suitable for widespread clinical implementation. We aimed to address this knowledge gap by systematically evaluating the performance of proposed prognostic models, among consecutive patients hospitalised for COVID-19 at a single centre.

## Methods

### Identification of candidate prognostic models

We used a published living systematic review to identify candidate prognostic models for COVID-19 indexed in PubMed, Embase, Arxiv, medRxiv, or bioRxiv until 5^th^May 2020^8^. We included models that aim to predict clinical deterioration or mortality among patients with COVID-19. We also included prognostic scores commonly used in clinical practice^10–12^, but not specifically developed for COVID-19 patients. For each candidate model identified, we extracted predictor variables, outcome definitions (including time horizons), modelling approaches, and final model parameters from original publications, and contacted authors for additional information where required. We excluded scores where the underlying model parameters were not publicly available, since we were unable to reconstruct them, along with models for which included predictors were not available in our dataset. The latter included models that require computed tomography imaging or arterial blood gas sampling, since these investigations were not routinely performed among unselected patients with COVID-19 at our centre.

### Study population

Our study is reported in accordance with transparent reporting of a multivariable prediction model for individual prognosis or diagnosis (TRIPOD) guidance for external validation studies^13^. We included consecutive adults admitted to University College Hospital London with a final diagnosis of PCR-confirmed or clinically diagnosed COVID-19, between 1^st^ February and 30^th^ April 2020. Since we sought to use data from the point of hospital admission to predict outcomes, we excluded patients transferred in from other hospitals, and those with hospital-acquired COVID-19 (defined as 1^st^ PCR swab sent >5 days from date of hospital admission, as a proxy for the onset of clinical suspicion of SARS-CoV-2 infection).

### Data sources and variables of interest

Data were collected by direct extraction from electronic health records, complemented by manual curation. Variables of interest in the dataset included: demographics (age, gender, ethnicity), comorbidities, clinical observations, laboratory measurements, radiology reports, and clinical outcomes. We defined ‘clinical deterioration’ as a composite outcome including initiation of ventilatory support (continuous positive airway pressure, non-invasive ventilation, high flow nasal cannula oxygen, invasive mechanical ventilation or extra-corporeal membrane oxygenation) or death, equivalent to World Health Organization Clinical Progression Scale ≥ 6^14^. The rationale for this composite outcome is to make the endpoint more generalisable between centres, since hospital respiratory management algorithms may vary substantially. Each chest radiograph was reported by a single radiologist, reflecting routine clinical conditions, using British Society of Thoracic Imaging criteria, and using a modified version of the Radiographic Assessment of Lung Edema (RALE) score^15,16^. Participants were followed-up clinically to the point of discharge from hospital. We extended follow-up beyond discharge by cross-checking NHS spine records to identify reported deaths post-discharge, thus ensuring >30 days’ follow-up for all participants.

### Statistical analyses

For each prognostic model included in the analyses, we reconstructed the model according to authors’ original descriptions, and sought to evaluate the model discrimination and calibration performance against their original intended endpoint. For models that provide online risk calculator tools, we validated our reconstructed models against original authors’ models, by cross-checking our predictions against those generated by the web-based tools for a random subset of participants.

For models that used ICU admission or death, or ‘severe’ COVID-19 or death, as composite endpoints, we used our ‘clinical deterioration’ endpoint as the primary outcome, as defined above. Where models specified their intended time horizon in their original description, we used this timepoint in the primary analysis, in order to ensure unbiased assessment of model calibration. Where the intended time horizon was not specified, we assessed the model to predict in-hospital deterioration or mortality, as appropriate.

For all models, we assessed discrimination by quantifying the area under the receiver operating characteristic curve (AUROC)^17^. For models that provided outcome probability scores, we assessed calibration by visualising calibration of predicted *vs*. observed risk using loess-smoothed and quartile plots, and by quantifying calibration slopes and calibration-in-the-large (CITL). A perfect calibration slope should be 1; slopes <1 indicate overfitting, while slopes >1 reflect underfitting. Ideal CITL is 0; CITL>0 indicates that predictions are systematically too low, while CITL<0 indicates that predictions are too high. For models with points-based scores, we assessed calibration visually by plotting model scores *vs*. actual outcome proportions. For models that provide probability estimates, but where the model intercept was not available, we calibrated the model to our dataset by calculating the intercept when using the model linear predictor as an offset term, leading to perfect CITL. This approach, by definition, overestimated calibration with respect to CITL, but allowed us to examine the calibration slope in our dataset.

We also assessed the discrimination of each candidate model for standardised outcomes of: (a) our composite endpoint of clinical deterioration; and (b) mortality, across a range of pre-specified time horizons from admission (7 days, 14 days, 30 days and any time during hospital admission), by calculating time-dependent AUROCs (with cumulative sensitivity and dynamic specificity)^18^. The rationale for this analysis was to harmonise endpoints, in order to facilitate more direct comparisons of discrimination between the candidate models.

In order to further benchmark the performance of candidate prognostic models, we then computed AUROCs for a limited number of univariable predictors considered to be of highest importance *a priori*, based on clinical knowledge and existing data, for prediction of our composite endpoints of clinical deterioration and mortality (7 days, 14 days, 30 days and any time during hospital admission). The *a priori* predictors of interest examined in this analysis were age, clinical frailty scale, oxygen saturation at presentation on room air, C-reactive protein and absolute lymphocyte count^8,19^.

We performed decision curve analyses to quantify the net benefit achieved by each model for predicting the intended endpoint, in order to inform clinical decision making across a range of risk:benefit ratios for an intervention or ‘treatment’^20^. In this approach, the risk:benefit ratio is analogous to the cut point for a statistical model above which the intervention would be considered beneficial (deemed the ‘threshold probability’). Net benefit was calculated as sensitivity × prevalence – (1 – specificity) × (1 – prevalence) × w where w is the odds at the threshold probability and the prevalence is the proportion of patients who experienced the outcome^20^. We calculated net benefit across a range of clinically relevant threshold probabilities, ranging from 0 to 0.5, since the risk:benefit ratio may vary for any given intervention (or ‘treatment’). We compared the utility of each candidate model against strategies of treating all and no patients, and against the best performing univariable predictor for in-hospital clinical deterioration, or mortality, as appropriate. We calculated ‘delta’ net benefit as net benefit when using the index model minus net benefit when: (a) treating all patients; and (b) using most discriminating univariable predictor. Decision curve analyses were done using the *rmda* package in R^21^.

We handled missing data using multiple imputation by chained equations^22^, using the *mice* package in R^23^. All variables in the final prognostic models were included in the imputation model to ensure compatibility^22^. A total of 10 imputed datasets were generated; discrimination and calibration metrics were pooled using Rubin’s rules^24^. Individual predictions for each prognostic model were averaged across imputations for each participant in order to generate pooled calibration plots, ROC curves and decision curves.

All analyses were conducted in R (version 3.5.1).

### Sensitivity analyses

We recalculated discrimination and calibration parameters for each candidate model using a complete case analysis. We also examined for non-linearity in the *a priori* univariable predictors using restricted cubic splines, with 3 knots. Finally, we estimated optimism for discrimination and calibration parameters for the *a priori* univariable predictors using bootstrapping (1,000 iterations), using the *rms* package in R^25^.

## Results

### Summary of candidate prognostic models

We identified a total of 37 studies describing prognostic models, of which 19 studies (including 22 unique models) were eligible for inclusion (Supplementary Figure 1 and Table 1). Of these, 5 models were not specific to COVID-19, but were developed as prognostic scores for emergency department attendees^26^, hospitalised patients^12,27^, people with suspected infection^10^ or community-acquired pneumonia^11^, respectively. Of the 17 models developed specifically for COVID-19, most (10/17) were developed using datasets originating in China. A total of 13/22 models use points-based scoring systems to derive final model scores, with the remainder using logistic regression modelling approaches to derive probability estimates. A total of 12/22 prognostic models primarily aimed to predict clinical deterioration, while the remaining 10 sought to predict mortality alone. When specified, time horizons for prognosis ranged from 1 to 30 days.

**Table 1:**
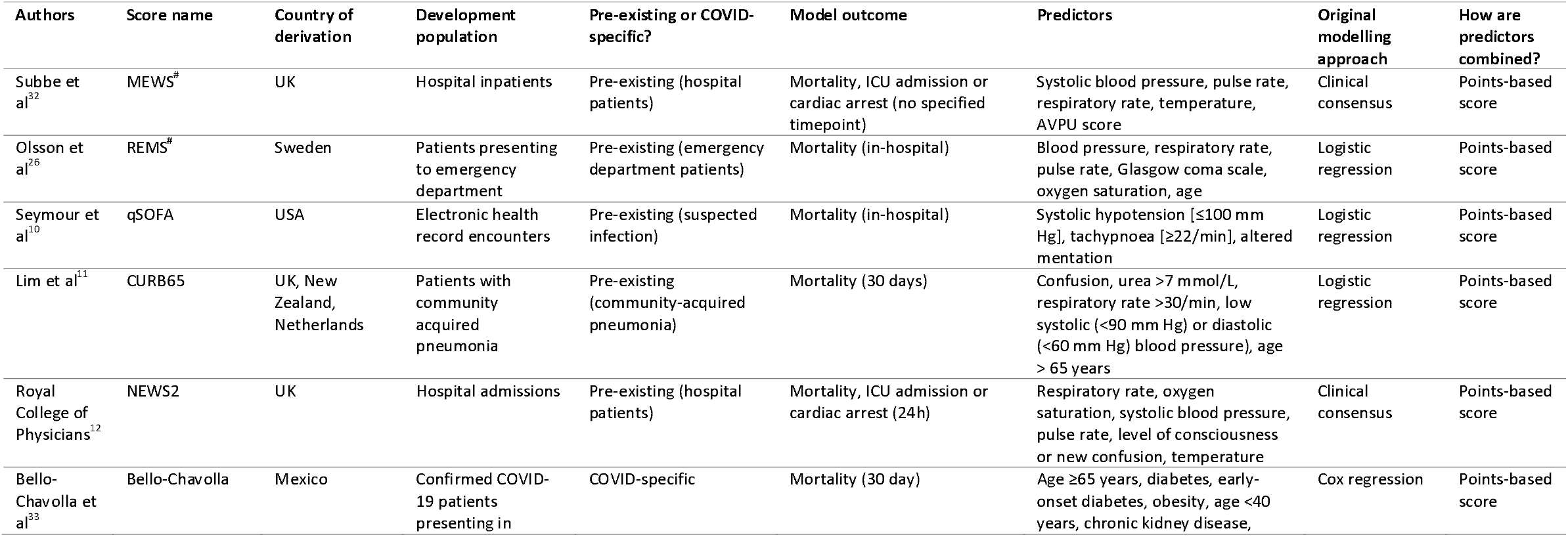
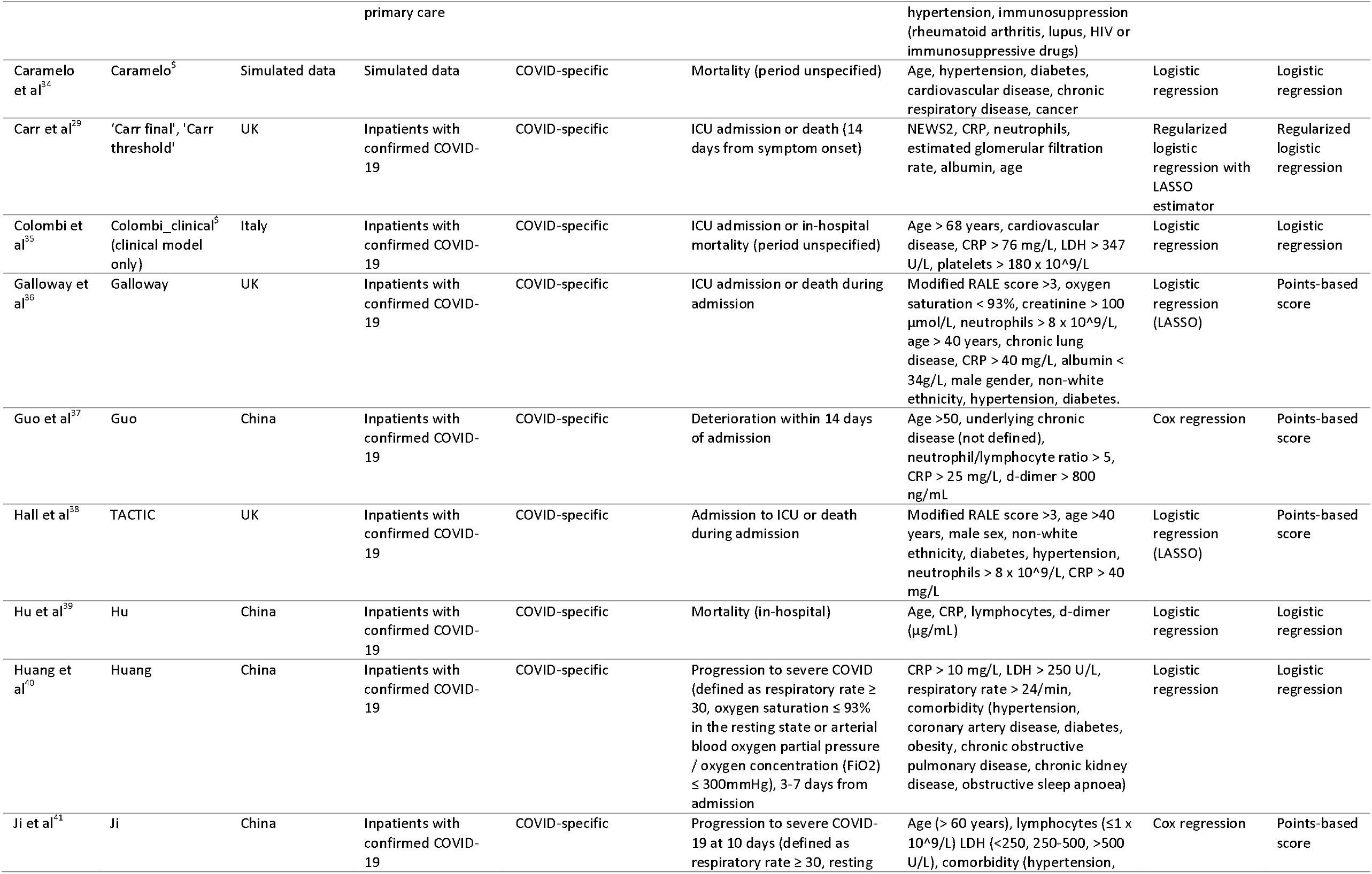
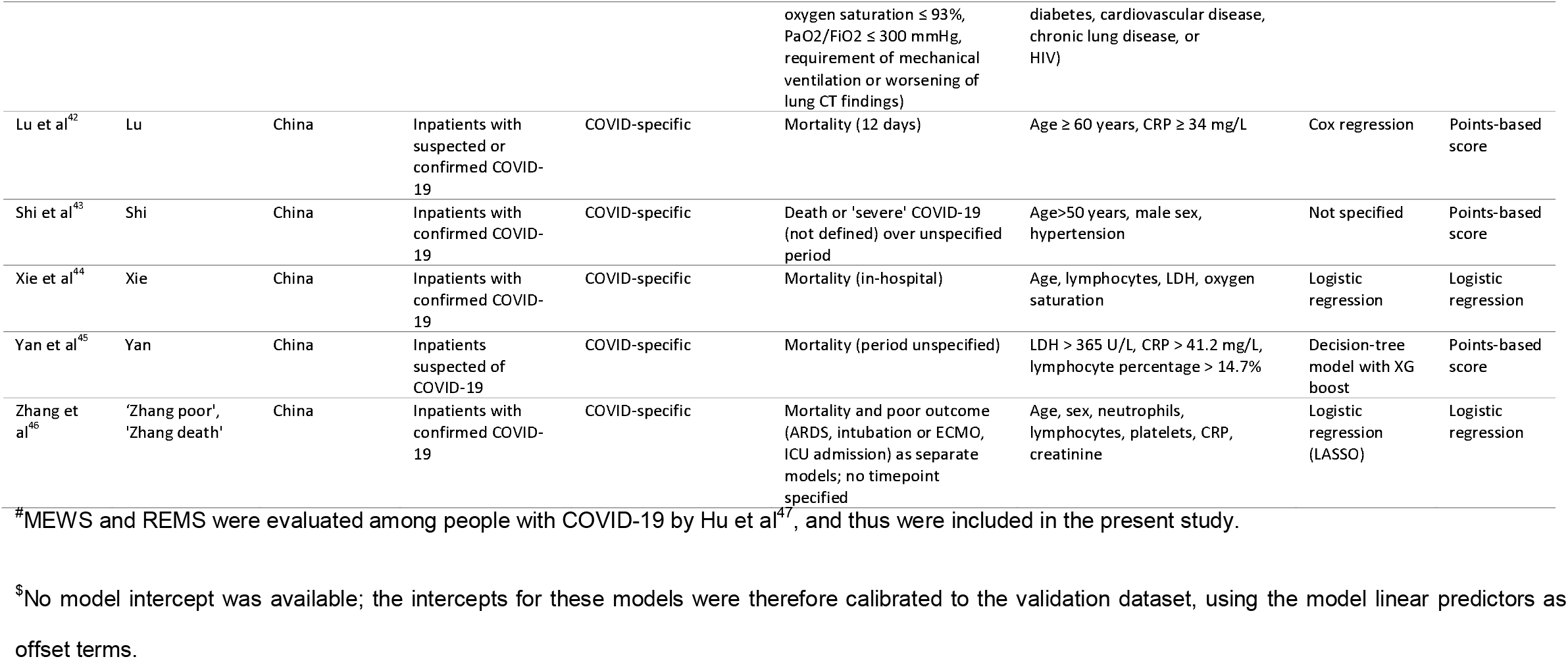
Characteristics of studies describing prognostic models included in systematic evaluation. MEWS = modified early warning score; qSOFA = quick sequential (sepsis-related) organ failure assessment; REMS = rapid emergency medicine score; NEWS = national early warning score; TACTIC = therapeutic study in pre-ICU patients admitted with COVID-19; AVPU = Alert / responds to voice / responsive to pain / unresponsive; CRP = C-reactive protein; LDH = lactate dehydrogenase; RALE = radiographic assessment of lung edema; ARDS = acute respiratory distress syndrome; ICU = intensive care unit; ECMO = extra-corporeal membrane oxygenation. Units, unless otherwise specified, are: age in years; respiratory rate in breaths per minute; heart rate in beats per minute; blood pressure in mmHg; temperature in °C; oxygen saturation in %; CRP in mg/L; LDH in U/L; neutrophils, lymphocytes, total white cell count and platelets × 10^9/L; D-dimer in ng/mL; creatinine in μmol/L; estimated glomerular filtration rate in mL/min/1.73 m2, albumin in g/L.

### Overview of study cohort

During the study period, 521 adults were admitted with a final diagnosis of COVID-19, of whom 411 met the eligibility criteria for inclusion (Supplementary Figure 2). Median age of the cohort was 66 years (interquartile range (IQR) 53-79), and the majority were male (252/411; 61.3%). Table 2 shows the baseline demographics, comorbidities, laboratory results and clinical measurements of the study cohort, of whom most (370/411; 90.0%) had PCR-confirmed SARS-CoV-2 infection. A total of 180 (43.8%) and 115 (28.0%) of participants met the endpoints of clinical deterioration and mortality, respectively, above the minimum requirement of 100 events recommended for external validation studies ^28^. The risks of clinical deterioration and death declined with time since admission (median days to deterioration 1.4 (IQR 0.3-4.2); median days to death 6.6 (IQR 3.6-13.1); Supplementary Figure 3). Most variables required for calculation of the 22 prognostic model scores were available among the vast majority of participants. However, admission lactate dehydrogenase was only available for 183/411 (44.5%) and D-dimer measured for 153/411 (37.2%). Supplementary Figure 4 shows missingness of each prognostic model in the complete case dataset, stratified by the outcomes of interest, due to unavailability of predictor variables.

**Table 2:**
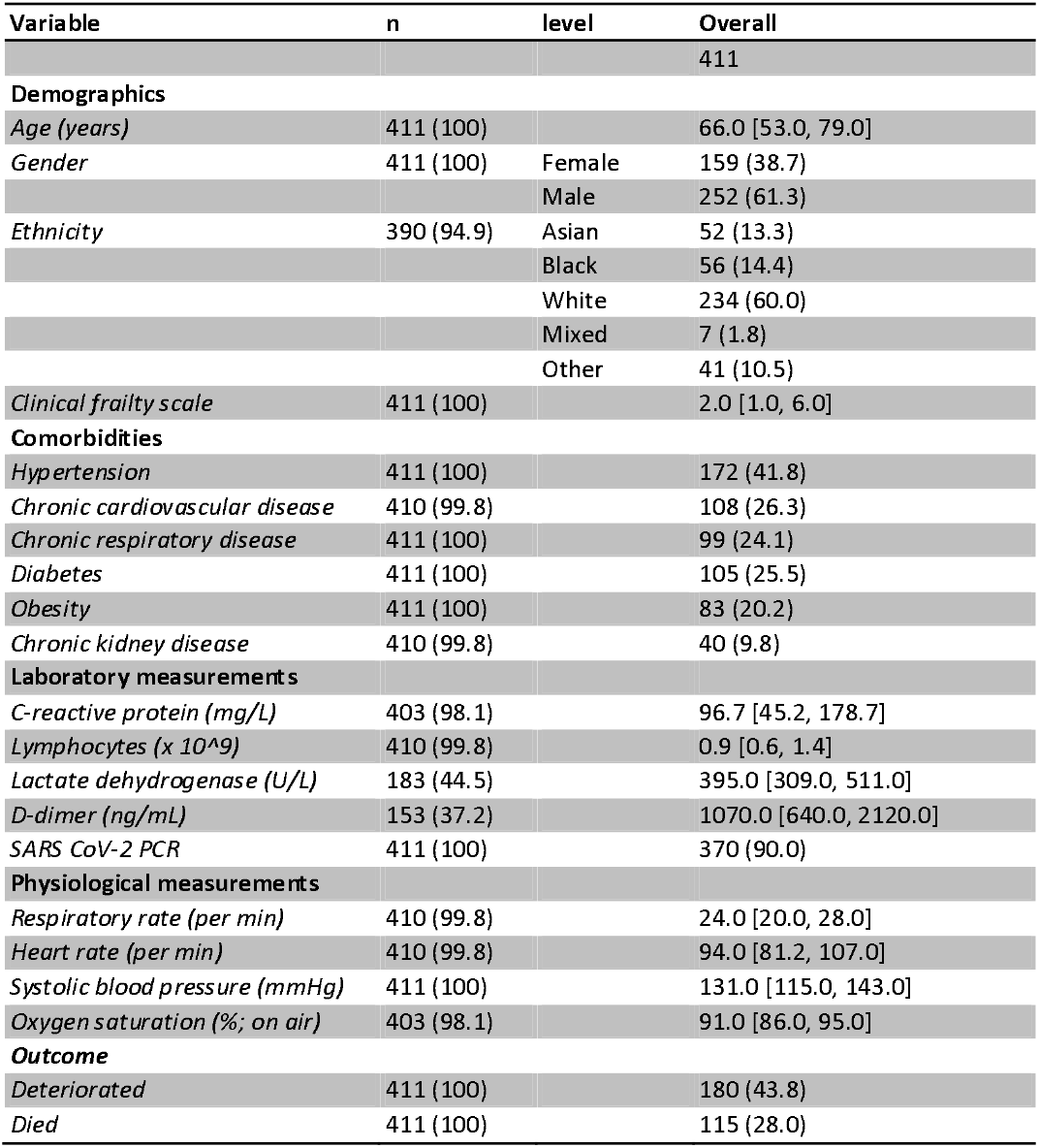
Baseline characteristics of hospitalised adults with COVID-19 included in systematic evaluation cohort. Laboratory and physiological measurements reflect parameters at the time of hospital admission. N column shows number of participants with available data for each variable. Data are shown as N (%) for categorical data or median (interquartile range (IQR)) for continuous variables.

| Variable | n | level | Overall |
| --- | --- | --- | --- |
|  |  |  | 411 |
| **Demographics** |  |  |  |
| Age (years) | 411 (100) |  | 66.0 [53.0, 79.0] |
| Gender | 411 (100) | Female | 159 (38.7) |
|  |  | Male | 252 (61.3) |
| Ethnicity | 390 (94.9) | Asian | 52 (13.3) |
|  |  | Black | 56 (14.4) |
|  |  | White | 234 (60.0) |
|  |  | Mixed | 7 (1.8) |
|  |  | Other | 41 (10.5) |
| Clinical frailty scale | 411 (100) |  | 2.0 [1.0, 6.0] |
| **Comorbidities** |  |  |  |
| Hypertension | 411 (100) |  | 172 (41.8) |
| Chronic cardiovascular disease | 410 (99.8) |  | 108 (26.3) |
| Chronic respiratory disease | 411 (100) |  | 99 (24.1) |
| Diabetes | 411 (100) |  | 105 (25.5) |
| Obesity | 411 (100) |  | 83 (20.2) |
| Chronic kidney disease | 410 (99.8) |  | 40 (9.8) |
| **Laboratory measurements** |  |  |  |
| C-reactive protein (mg/L) | 403 (98.1) |  | 96.7 [45.2, 178.7] |
| Lymphocytes ($\times 10^9$) | 410 (99.8) | | 0.9 [0.6, 1.4] |
| Lactate dehydrogenase (U/L) | 183 (44.5) |  | 395.0 [309.0, 511.0] |
| D-dimer (ng/mL) | 153 (37.2) |  | 1070.0 [640.0, 2120.0] |
| SARS CoV-2 PCR | 411 (100) |  | 370 (90.0) |
| **Physiological measurements** |  |  |  |
| Respiratory rate (per min) | 410 (99.8) |  | 24.0 [20.0, 28.0] |
| Heart rate (per min) | 410 (99.8) |  | 94.0 [81.2, 107.0] |
| Systolic blood pressure (mmHg) | 411 (100) |  | 131.0 [115.0, 143.0] |
| Oxygen saturation (%; on air) | 403 (98.1) |  | 91.0 [86.0, 95.0] |
| **Outcome** |  |  |  |
| Deteriorated | 411 (100) |  | 180 (43.8) |
| Died | 411 (100) |  | 115 (28.0) |

### Evaluation of prognostic models for original primary outcomes

Table 3 shows discrimination and calibration metrics, where appropriate, for the 22 evaluated prognostic models in the primary multiple imputation analysis. The highest AUROCs were achieved by the NEWS2 score for prediction of deterioration over 24 hours (0.78; 95% CI 0.73 - 0.83), and the Carr ‘final’ model for prediction of deterioration over 14 days (0.78; 95% CI 0.74 - 0.82). Of the other prognostic scores currently used in routine clinical practice, CURB65 was noted to have reasonable discrimination for 30-day mortality (AUROC 0.75; 95% CI 0.70 - 0.80), and qSOFA discriminated in-hospital mortality poorly (AUROC 0.6; 95% CI 0.55 - 0.65). ROC curves are shown for each candidate model in Supplementary Figure 5.

**Table 3:**
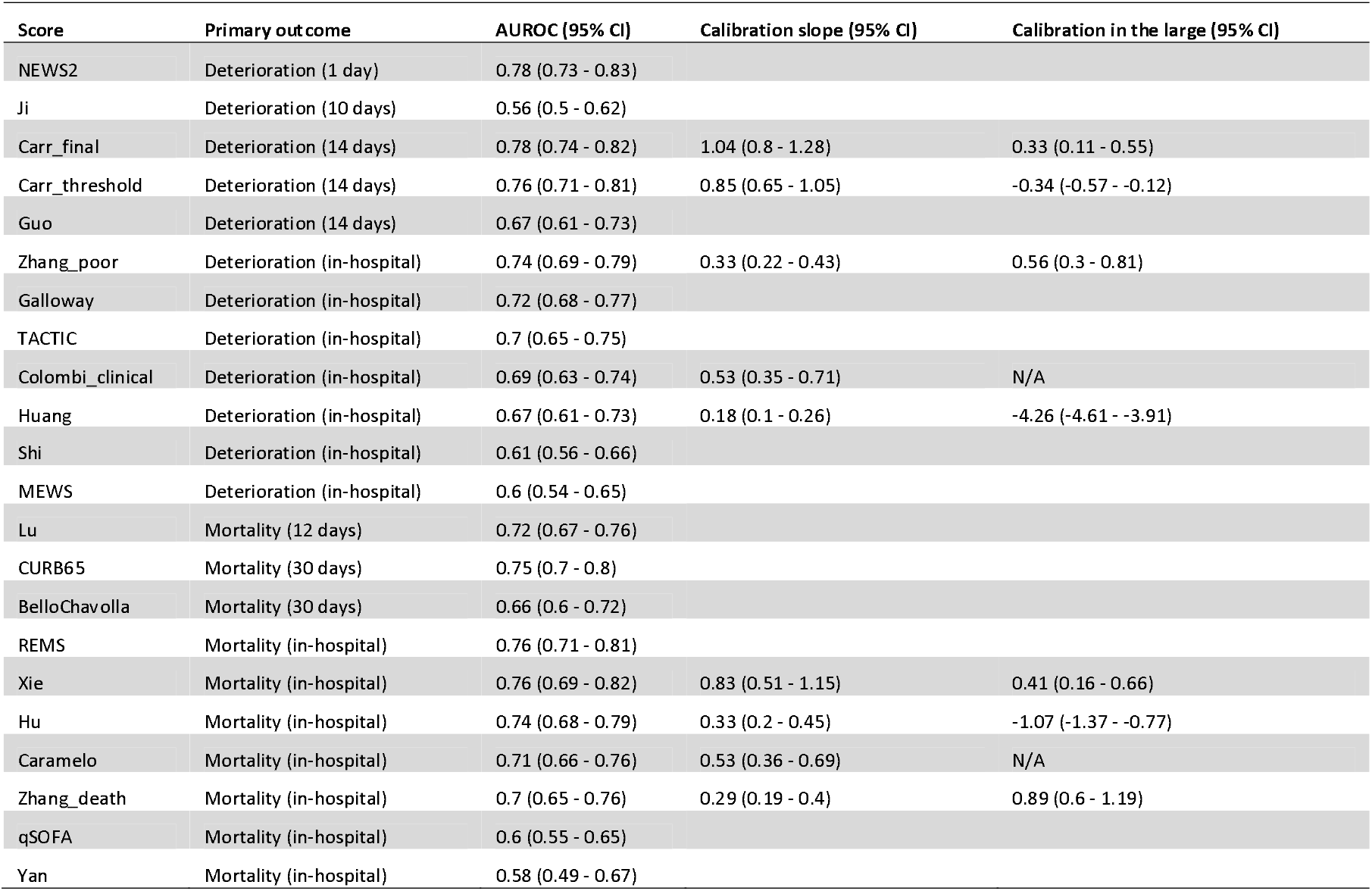
Validation metrics of prognostic scores for COVID-19, using primary multiple imputation analysis (n=411). For each model, performance is evaluated for its original intended outcome, shown in ‘Primary outcome’ column. AUROC = area under the receiver operating characteristic curve; CI = confidence interval.

| Score | Primary outcome | AUROC (95% CI) | Calibration slope (95% CI) | Calibration in the large (95% CI) |
| --- | --- | --- | --- | --- |
| NEWS2 | Deterioration (1 day) | 0.78 (0.73 - 0.83) |  |  |
| Ji | Deterioration (10 days) | 0.56 (0.5 - 0.62) |  |  |
| Carr_final | Deterioration (14 days) | 0.78 (0.74 - 0.82) | 1.04 (0.8 - 1.28) | 0.33 (0.11 - 0.55) |
| Carr_threshold | Deterioration (14 days) | 0.76 (0.71 - 0.81) | 0.85 (0.65 - 1.05) | -0.34 (-0.57 - -0.12) |
| Guo | Deterioration (14 days) | 0.67 (0.61 - 0.73) |  |  |
| Zhang_poor | Deterioration (in-hospital) | 0.74 (0.69 - 0.79) | 0.33 (0.22 - 0.43) | 0.56 (0.3 - 0.81) |
| Galloway | Deterioration (in-hospital) | 0.72 (0.68 - 0.77) |  |  |
| TACTIC | Deterioration (in-hospital) | 0.7 (0.65 - 0.75) |  |  |
| Colombi_clinical | Deterioration (in-hospital) | 0.69 (0.63 - 0.74) | 0.53 (0.35 - 0.71) | N/A |
| Huang | Deterioration (in-hospital) | 0.67 (0.61 - 0.73) | 0.18 (0.1 - 0.26) | -4.26 (-4.61 - -3.91) |
| Shi | Deterioration (in-hospital) | 0.61 (0.56 - 0.66) |  |  |
| MEWS | Deterioration (in-hospital) | 0.6 (0.54 - 0.65) |  |  |
| Lu | Mortality (12 days) | 0.72 (0.67 - 0.76) |  |  |
| CURB65 | Mortality (30 days) | 0.75 (0.7 - 0.8) |  |  |
| BelloChavolla | Mortality (30 days) | 0.66 (0.6 - 0.72) |  |  |
| REMS | Mortality (in-hospital) | 0.76 (0.71 - 0.81) |  |  |
| Xie | Mortality (in-hospital) | 0.76 (0.69 - 0.82) | 0.83 (0.51 - 1.15) | 0.41 (0.16 - 0.66) |
| Hu | Mortality (in-hospital) | 0.74 (0.68 - 0.79) | 0.33 (0.2 - 0.45) | -1.07 (-1.37 - -0.77) |
| Caramelo | Mortality (in-hospital) | 0.71 (0.66 - 0.76) | 0.53 (0.36 - 0.69) | N/A |
| Zhang_death | Mortality (in-hospital) | 0.7 (0.65 - 0.76) | 0.29 (0.19 - 0.4) | 0.89 (0.6 - 1.19) |
| qSOFA | Mortality (in-hospital) | 0.6 (0.55 - 0.65) |  |  |
| Yan | Mortality (in-hospital) | 0.58 (0.49 - 0.67) |  |  |

For all models that provide probability scores for either deterioration or mortality, calibration appeared visually poor with evidence of overfitting and either systematic overestimation or underestimation of risk (Figure 1). Supplementary Figure 6 shows associations between prognostic models with points-based scores and actual risk. In addition to demonstrating reasonable discrimination, the NEWS2 and CURB65 models demonstrated approximately linear associations between scores and actual probability of deterioration at 24 hours and mortality at 30 days, respectively.

**Figure 1:**
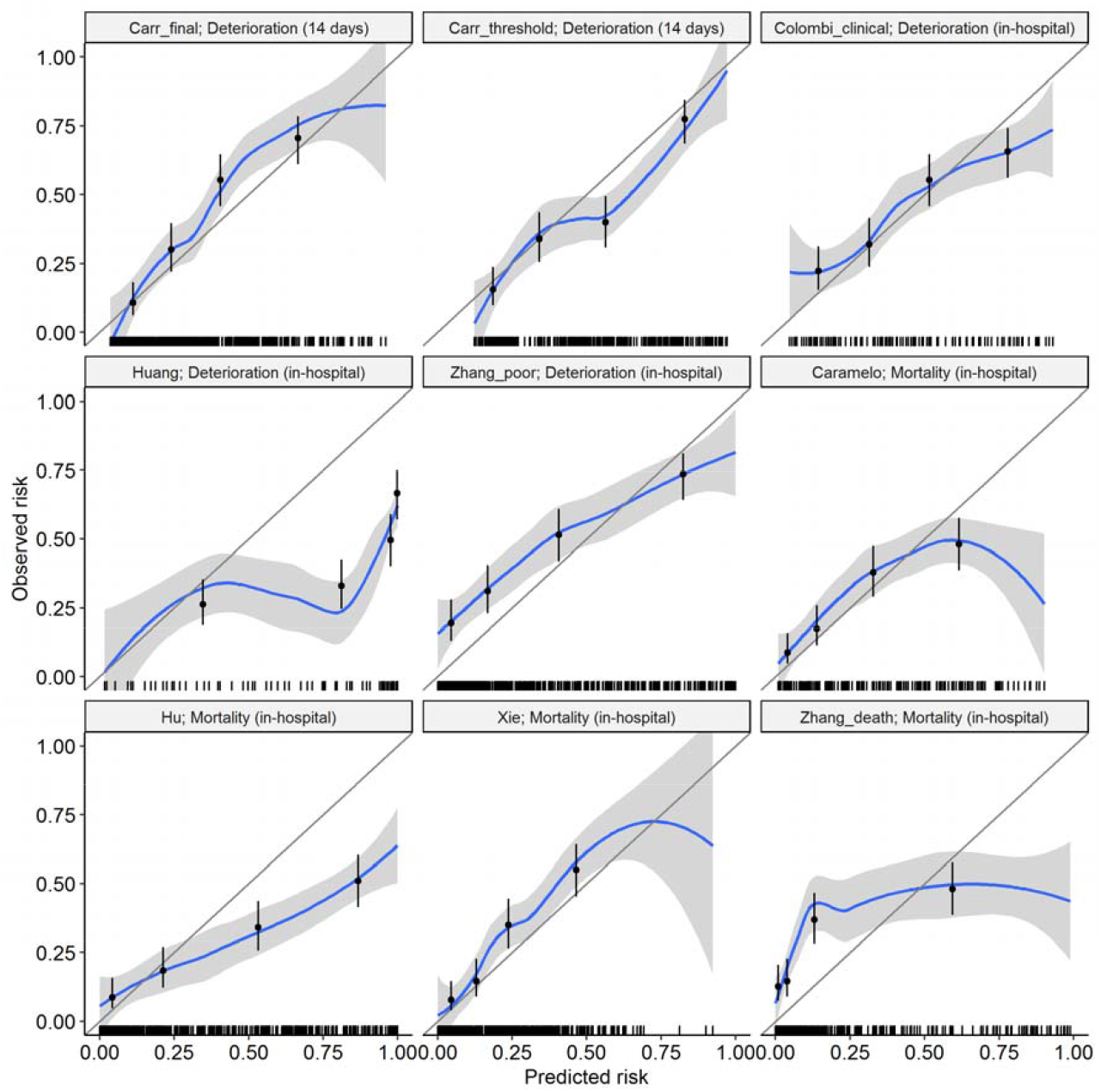
Calibration plots for prognostic models estimating outcome probabilities. For each plot, the blue line represents a Loess-smoothed calibration curve, and scatter points show quartiles of predicted risk. Rug plots indicate the distribution of data points. No model intercept was available for the Caramelo or Colombi ‘clinical’ models; the intercepts for these models were calibrated to the validation dataset, by using the model linear predictors as offset terms. Calibration-in-the-large is therefore not shown for these models, since it is zero by definition. The primary outcome of interest for each model is shown in the plot sub-heading. Individual predictions for each prognostic model were averaged across imputations for each participant in the dataset in order to generate these pooled calibration plots.

### Time-dependent discrimination of candidate models and a priori univariable predictors for standardised outcomes

Next, we sought to compare the discrimination of these models for different outcomes across the range of time horizons, benchmarked against preselected univariable predictors associated with adverse outcomes in COVID-19^8,19^. We recalculated time-dependent AUROCs for each of these outcomes, stratified by time horizon to the outcome (Supplementary Figures 7 and 8). These analyses showed that AUROCs generally declined with increasing time horizons. Admission oxygen saturation on room air was the strongest predictor of in-hospital deterioration (AUROC 0.76; 95% CI 0.71-0.81), while age was the strongest predictor of in-hospital mortality (AUROC 0.76; 95% CI 0.71-0.81).

### Decision curve analyses to assess clinical utility

We compared net benefit for each prognostic model (for its original intended endpoint) to the strategies of treating all patients, treating no patients, and using the most discriminating univariable predictor for either deterioration (i.e. oxygen saturation on air) or mortality (i.e. patient age) to stratify treatment (Supplementary Figure 9). Although all prognostic models showed greater net benefit than treating all patients at the higher range of threshold probabilities, none of these models demonstrated consistently greater net benefit than the most discriminating univariable predictor, across the range of threshold probabilities (Figure 2).

**Figure 2:**
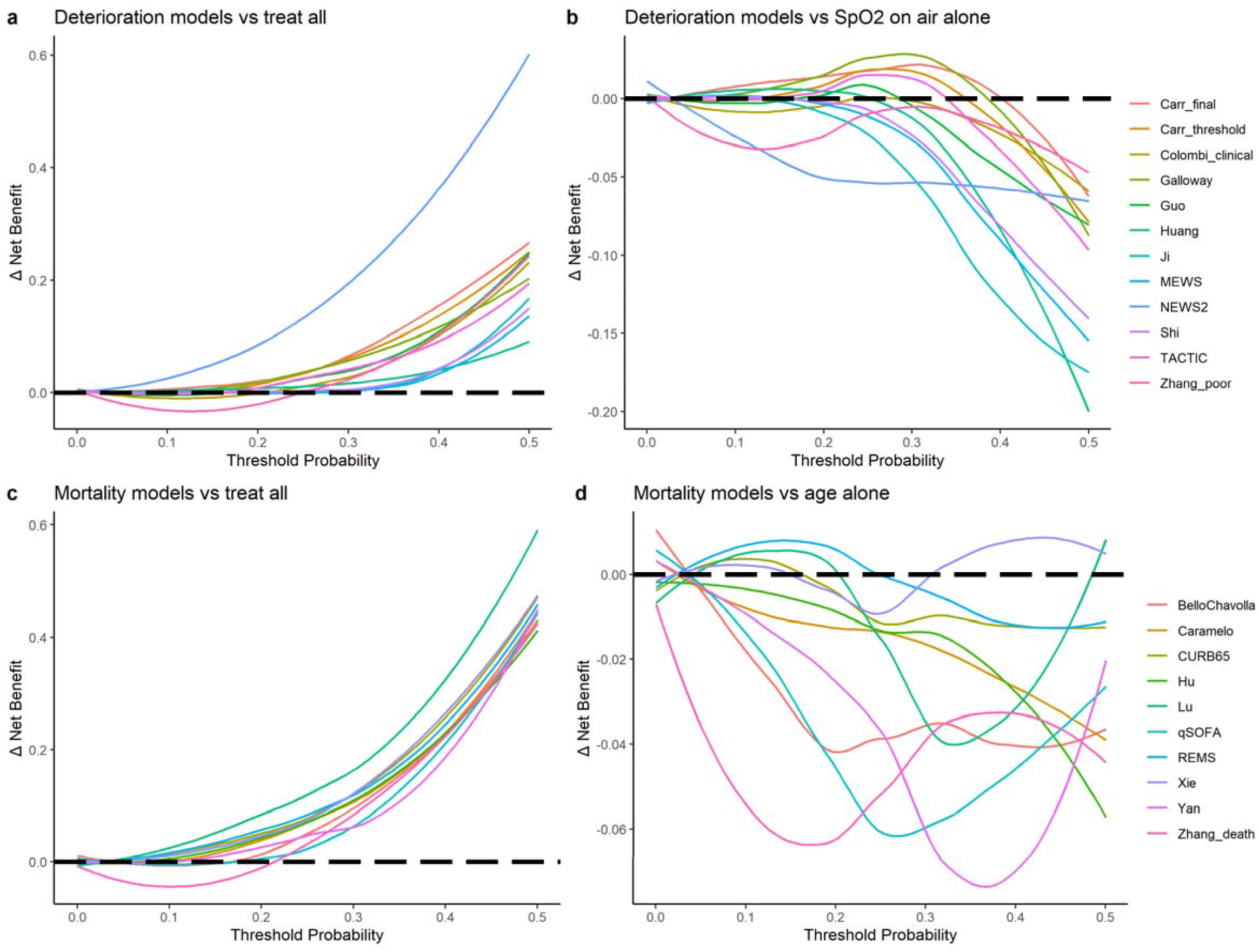
Decision curve analysis showing delta net benefit of each candidate model, compared to treating all patients and best univariable predictors. For each analysis, the endpoint is the original intended outcome and time horizon for the index model. Delta net benefit is calculated as net benefit when using the index model minus net benefit when: (1) treating all patients; and (2) using the most discriminating univariable predictor. The most discriminating univariable predictor is admission oxygen saturation (SpO2) on room air for deterioration models and patient age for mortality models. Individual predictions for each prognostic model were averaged across imputations for each participant in the dataset in order to generate pooled decision curve plots. Delta net benefit is shown with Loess-smoothing. Black dashed line indicates threshold above which index model has greater net benefit than the comparator. Full decision curves for each candidate model are shown in Supplementary Figure 9.

### Sensitivity analyses

Recalculation of model discrimination and calibration metrics for prediction of the original intended endpoint using a complete case analysis revealed similar results to the primary multiple imputation approach (Supplementary Table 1). Visual examination of associations between the most discriminating univariable predictors and log odds of deterioration or death using restricted cubic splines showed no evidence of non-linear associations (Supplementary Figure 10). Finally, internal validation using bootstrapping showed near zero optimism for discrimination and calibration parameters for the univariable models (Supplementary Table 2).

## Discussion

In this observational cohort study of consecutive adults hospitalised with COVID-19, we systematically evaluated the performance of 22 prognostic models for COVID-19. These included models developed specifically for COVID-19, along with existing scores in routine clinical use prior to the pandemic. For prediction of both clinical deterioration or mortality, discrimination appeared modest or poor for most models. NEWS2 performed reasonably well for prediction of deterioration over a 24-hour interval, achieving an AUROC of 0.78, while the Carr ‘final’ model^29^ also had reasonable discrimination (AUROC 0.78), but tended to systematically underestimate risk. All COVID-specific models that derived an outcome probability of either deterioration or mortality showed poor calibration. We found that oxygen saturation (AUROC 0.76) and patient age (AUROC 0.76) were the most discriminating single variables for prediction of in-hospital deterioration and mortality respectively. These predictors have the added advantage that they are immediately available at the point of presentation to hospital. In decision curve analysis, no prognostic model demonstrated clinical utility consistently greater than using oxygen saturation on room air to predict deterioration, or patient age to predict mortality.

While previous studies have largely focused on novel model discovery, or evaluation of a limited number of existing models, this is the first study to our knowledge to evaluate systematically-identified candidate prognostic models for COVID-19. We used a comprehensive living systematic review^8^ to identify eligible models and sought to reconstruct each model as per the original authors’ description. We then evaluated performance against its intended outcome and time horizon, wherever possible, using recommended methods of external validation incorporating assessments of discrimination, calibration and net benefit^17^. Moreover, we used a robust approach of electronic health record data capture, supported by manual curation, in order to ensure a high-quality dataset, and inclusion of unselected and consecutive COVID-19 cases that met our eligibility criteria. In addition, we used robust outcome measures of mortality and clinical deterioration, aligning with the WHO Clinical Progression Scale^14^

A weakness of the current study is that it is based on data from a single centre, and therefore cannot assess between-setting heterogeneity in model performance. Second, due to the limitations of routinely collected data, predictor variables were available for varying numbers of participants for each model. We therefore performed multiple imputation, in keeping with recommendations for development and validation of multivariable prediction models, in our primary analyses^30^. Findings were similar in the complete case sensitivity analysis, thus supporting the robustness of our results. Thirdly, a number of models could not be reconstructed in our data. For some models, this was due the absence of predictors in our dataset, such as those requiring computed tomography imaging, since this is not currently routinely recommended for patients with suspected or confirmed COVID-19^16^. We were also not able to include models for which the parameters were not publicly available. This underscores the need for strict adherence to reporting standards in multivariable prediction models^13^. Finally, we used admission data only as predictors in this study, since most prognostic scores are intended to predict outcomes at the point of hospital admission. We note, however, that some scores (such as NEWS2) are designed for dynamic in-patient monitoring. Future studies may integrate serial data to examine model performance when using such dynamic measurements.

Despite the vast global interest in the pursuit of prognostic models for COVID-19, our findings show that no COVID-19-specific models can currently be recommended for routine clinical use. All novel prognostic models for COVID-19 assessed in the current study were derived from single-centre data. Future studies may seek to pool data from multiple centres in order to robustly evaluate the performance of existing models across heterogeneous populations, and develop and validate novel prognostic models, through individual participant data meta-analysis^31^. Such an approach would allow assessments of between-study heterogeneity and the likely generalisability of candidate models. It is also imperative that discovery populations are representative of target populations for model implementation, with inclusion of unselected cohorts. Moreover, we strongly advocate for transparent reporting in keeping with TRIPOD standards (including modelling approaches, all coefficients and standard errors) along with standardisation of outcomes and time horizons, in order to facilitate ongoing systematic evaluations of model performance and clinical utility^13^.

We conclude that baseline oxygen saturation on room air and patient age are strong predictors of deterioration and mortality, respectively. None of the prognostic models evaluated in this study offer incremental value for patient stratification to these univariable predictors. Therefore, none of the evaluated prognostic models for COVID-19 can be recommended for routine clinical implementation. Future studies seeking to develop prognostic models for COVID-19 should consider integrating multi-centre data in order to increase generalisability of findings, and should ensure benchmarking against existing models and simpler univariable predictors.

## Data Availability

The conditions of regulatory approvals for the present study preclude open access data sharing to minimise risk of patient identification through granular individual health record data. The authors will consider specific requests for data sharing as part of academic collaborations subject to ethical approval and data transfer agreements in accordance with GDPR regulations.

## Footnotes

## Acknowledgements

The UCLH COVID-19 Reporting Group was comprised of the following individuals, who were involved in data curation as non-author contributors: Asia Ahmed, Ronan Astin, Malcolm Avari, Elkie Benhur, Anisha Bhagwanani, Timothy Bonnici, Sean Carlson, Jessica Carter, Sonya Crowe, Mark Duncan, Ferran Espuny-Pujol, James Fullerton, Marc George, Georgina Harridge, Ali Hosin, Rachel Hubbard, Adnan Hubraq, Prem Jareonsettasin, Zella King, Avi Korman, Sophie Kristina, Lawrence Langley, Jacques-Henri Meurgey, Henrietta Mills, Alfio Missaglia, Ankita Mondal, Samuel Moulding, Christina Pagel, Liyang Pan, Shivani Patel, Valeria Pintar, Jordan Poulos, Ruth Prendecki, Alexander Procter, Magali Taylor, David Thompson, Lucy Tiffen, Hannah Wright, Luke Wynne, Jason Yeung, Claudia Zeicu, Leilei Zhu

## Author contributions

RKG and MN conceived the study. RKG conducted the analysis and wrote the first draft of the manuscript. All other authors contributed towards data collection, study design and/or interpretation. All authors have critically appraised and approved the final manuscript prior to submission. The corresponding author attests that all listed authors meet authorship criteria and that no others meeting the criteria have been omitted.

Members of The UCLH COVID-19 Reporting contributed towards data curation and are non-author contributors/collaborators for this study.

## Ethical approval

This study was approved by East Midlands - Nottingham 2 Research Ethics Committee (REF: 20/EM/0114).

